# Charcot-Marie-Tooth disease type 1E: Clinical Natural History and Molecular Impact of *PMP22* Variants

**DOI:** 10.1101/2025.05.01.25326605

**Authors:** Kailee S. Ward, Christopher P. Ptak, Natalya Pashkova, Tiffany Grider, Tabitha A. Peterson, Davide Pareyson, Chiara Pisciotta, Paola Saveri, Isabella Moroni, Matilde Laura, Joshua Burns, Manoj P. Menezes, Kayla Cornett, Richard Finkel, Bipasha Mukherjee-Clavin, Charlotte J. Sumner, Maxwell Greene, Omer Abdul Hamid, David Herrmann, Reza Sadjadi, David Walk, Stephan Züchner, Mary M. Reilly, Steven S. Scherer, Inherited Neuropathy Consortium, Robert C. Piper, Michael E. Shy

## Abstract

Charcot-Marie-Tooth disease type 1E (CMT1E) is a rare, autosomal dominant peripheral neuropathy caused by missense variants, deletions, and truncations within the *peripheral myelin protein-22* (*PMP22*) gene. CMT1E phenotypes vary depending on the specific variant, ranging from mild to severe, and there is little natural history and phenotypic progression data on individuals with CMT1E. Patients with CMT1E were evaluated during initial and follow-up visits at sites within the Inherited Neuropathy Consortium. Clinical characteristics were obtained from history, neurological exams, and nerve conduction studies. Clinical outcome measures were used to quantify baseline and longitudinal changes, including the Rasch-modified CMT Examination Score version 2 (CMTESv2-R) and the CMT Pediatric Scale (CMTPedS). The trafficking of PMP22 variants in transfected cells was correlated to disease severity. Twenty-four, presumed disease-causing *PMP22* variants were identified in 50 individuals from 35 families, including 19 missense variants, three in-frame deletions, and two truncations. Twenty-nine patients presented with delayed walking during childhood. At their baseline evaluation, the mean CMTESv2-R in 46 patients was 16 ± 7.72 (out of 32), and the mean CMTPedS from 17 patients was 28 ± 6.35 (out of 44). Six individuals presented with hearing loss, eleven with scoliosis, three with hip dysplasia, and one with both scoliosis and hip dysplasia. Twenty variants were localized within in transmembrane domains; 31 of 35 individuals with these variants had moderate to severe phenotypes. Three variants were found in the extracellular domain and were associated with milder phenotypes. Reduced expression of PMP22 at the cell surface, and the location of missense variants within in the transmembrane domain correlated with disease severity. Pathogenic *PMP22* variants located within the transmembrane regions usually cause a moderate to severe clinical phenotype, beginning in early childhood, and have impaired trafficking to the plasma membrane.

## Introduction

Charcot-Marie-Tooth disease (CMT) refers to heritable peripheral neuropathies, which are a common genetic disease, affecting 1:2500 individuals ^1^. Autosomal dominant inheritance is the most common, followed by X-linked dominant and autosomal recessive types. In the demyelinating forms of CMT (CMT1 and CMT4), the expression of the variant allele(s) in myelinating Schwann cells causes a demyelinating neuropathy. In the axonal forms of CMT (CMT2 and others), the expression of the variant allele(s) in the neurons of the peripheral nervous system (PNS) causes an axonal neuropathy. CMT1A is the most common subtype, and is caused by a 1.5 Mb duplication within chromosome 17p11.2, resulting in an extra copy of *PMP22* gene ^2, 3^. Deletion of this same 1.5 Mb region results in a heterozygous deletion of *PMP22*, reducing PMP22 protein and causing a distinct neuropathy, Hereditary Neuropathy with Liability to Pressure Palsies (HNPP), that is milder than CMT1A ^4^. Finally, missense variants, deletions, and some truncations within the *PMP22* gene usually cause demyelinating CMT and are classified as CMT1E, with variable phenotypes that are more severe than HNPP ^5, 6^, indicating that they cause a toxic gain of function. The pathogenic mechanisms of the CMT1E variants are incompletely understood, and some variants appeared to be retained in the endoplasmic reticulum (ER)^7–10^. In contrast to CMT1A ^11^, natural history studies of CMT1E have not been performed; case reports describe severe, infantile-onset, CMT1A-like, and HNPP-like phenotypes ^5^.

PMP22 is a tetraspan protein expressed predominantly by myelinating Schwann cells as part of a coordinated program of myelin gene expression. It is localized to the compact myelin sheath, which is a multi-lamellar spiral of specialized plasma membrane that contains specialized lipids and proteins ^12, 13^ PMP22 is glycosylated at N41, initially in the ER and further modified in the Golgi apparatus ^14^. The majority of PMP22 undergoes rapid turnover, with only a smal fraction incorporating into compact myelin ^15^. Although the stability and even the proper formation of PNS myelin sheaths depends on the amount of PMP22 ^16, 17^, the precise function of PMP22 is not well understood. Reconstituted PMP22 and lipids form myelin-like assemblies ^18^. PMP22 forms a complex with itself, and some PMP22 variants promote self-aggregation ^10, 19–22^. The second transmembrane domain of PMP22 interacts with myelin protein zero (MPZ) ^23^, which also plays an essential role in the formation of compact PNS myelin ^24, 25^. To understand how *PMP22* variants cause CMT1E, we queried the registry of the Inherited Neuropathy Consortium (INC), which currently contains clinical data on over 8,500 individuals with CMT, including those with CMT1E. We identified 24, presumed disease-causing *PMP22* variants in 50 patients from 35 families, and found that disease severity correlated with the location of the CMT1E variant in the predicted structure of PMP22, and the extent to which the variants showed impaired trafficking to the plasma membrane.

## Methods and materials

### Study Design, Patient Recruitment, and Ethics

Patients included in this study were enrolled through the Inherited Neuropathy Consortium Rare Disease Clinical Research Network (INC RDCRN) 6601 natural history protocol (ClinicalTrials.gov, NCT01193075). Approval was obtained at all participating institutions from the institutional review boards and research ethics committees. Patients were enrolled and evaluated at one of 25 sites in the US, UK, Italy, and Australia between 2012 and 2024 and at Wayne State University between 2010 and 2012. Genetic testing was performed in all patients or in first- or second-degree relatives to confirm the *PMP22* sequence variant. All research participants and their guardians reviewed and signed the appropriate informed consent/assent forms. Demographic, genetic data and medical history were collected at the baseline assessment. Longitudinal follow-up data, including clinical presentation and examination, was collected prospectively on an annual basis. Functional research testing was performed on pediatric patients at baseline and prospectively during annual visits.

### *PMP22* Variant Curation

Variant classification in this study was completed according to the standards and guidelines outlined by the American College of Medical Genetics and Genomics and the Association for Molecular Pathology ^26, 27^. The inheritance pattern for these variants was determined by family history, clinical genetic testing, clinical examiniation, and electrophysiology of the proband’s parents when possible to classify a variant as *de novo* (Supplementary Fig. 1 and Supplementary Table 1). Affected individuals with no reported family history and untested parents were classified as sporadic. Pedigrees for common variants can be seen in Supplementary Fig. 2.

### *PMP22* Variant Depiction and Analysis

Using the PMP22 protein sequence (UniProt: Q01453), the variants were labelled on a 2-dimensional topology map. The variant amino acids substitutions were indicated by spheres at their Cα atom positions on a 3-dimensional model of PMP22 (AF-Q01453-F1-v4) obtained from the AlphaFold PSDB ^28^. The PPM method was used to define the transmembrane segments of PMP22 within a lipid bilayer and to generate a prediction for membrane depth ^29^. A similar transmembrane toplogy map for PMP22 has been described previously ^22^. The calculated membrane depth parameter used the average membrane depth of atoms in the amino acid and 0 Å as the depth at the water – lipid bilayer interface. The visualization of amino acid position specific data on the 3-dimensional PMP22 model was performed using UCSF Chimera ^30^. Whether amino acids were surface accessible was also calculated by UCSF Chimera. PMP22 deletion, truncation, and insertion models were generated using AlphaFold 3 ^31^.

### Clinical Outcome Measures

The clinical outcome measures included the CMT Examination Score version 2 (CMTESv2) ^32^ and the Rasch-weighted modified (CMTESv2-R), which has been shown to better predict disease progression in CMT1A ^11, 33^. The CMTESv2-R was collected at baseline and follow-up visits to measure clinical phenotypes throughout the patient’s lifespan. To link the genetic data with the associated phenotypes, all available CMTESv2-R scores were analyzed in relation to the age of the patient and the variant location within the PMP22 protein.

For patients aged 4-20, the CMT Pediatric Scale (CMTPedS) was administered, which is a validated performance-based outcome measure with 11 items covering strength, dexterity, sensation, gait, balance, power, and endurance ^34^. The CMTPedS total score (0-44, based on the 1000 Norms Project) was calculated at www.ClinicalOutcomeMeasures.org during baseline and follow-up visits to assess clinical phenotype presentation throughout childhood. All available CMTPedS scores were analyzed in concert with patient age and *PMP22* variant.

Higher CMTESv2-R and CMTPedS scores indicate an increased level of disease severity. Clinical investigators and assessors at each site were certified in these outcomes prior to use. Standard Response Means (SRM) were calculated to estimate the responsiveness of CMTES-R^11, 35^ and CMTPedS ^36^ over two year intervals. An SRM of 0.2-0.5 suggested low responsiveness, 0.5-0.8 moderate responsiveness and > 0.8 large responsiveness of the outcome measure.

### Curve Fitting and Statistical Analysis

Non-linear curve fitting of age-dependent progression was conducted for individual patient data on plots of CMTESv2 vs age using Origin7 and fit to sigmoidal Gompertz curves given by the general equation, y = a*exp(-exp(-k*(x-xc))). Age-adjusted CMTESv2-R values, b, were obtained by adjusting the age of center point, xc = 80 - 7.5*b, and the growth rate of the curve, k = 0.05*exp(0.1*b). The range for b = 0 to 10 approximately covers the range for patient CMTESv2-R scores. The amplitude, a, was fixed at 32. Statistical analysis of box and correlation plots was performed using R (v4.4.2) and RStudio (v2024.09.1).

### Cell Culture and Flow Cytometry Cell Surface Immunostaining

The missense variants listed in Supplementary Table 3 were made using a PMP22 expression plasmid encoding *PMP22* carrying a myc epitope within the 2^nd^ extracellular loop (PMP22-myc^exo^) (Supplementary Fig. 5B) and co-expressing a nuclear-localized monomeric Blue Fluorescent protein (mTAGBFP2) and the puromycin-resistance protein as previously described ^23^. This plasmid flanks the genes of interest with insulators for durable expression and sequences for piggyBac-mediated insertion into the genome ^37^. The plasmids for PMP22-GFP and PMP22myc^exo^ fusion protein expression were made as described ^23^. The plasmid for mTAGBFP-KDEL expression was made by substituting mTAGBFP in a previously described plasmid encoding GFP-KDEL^38^. HEK293 cells *(ATCC: CRL-1573)* were cultured in Dulbecco’s modified Eagle’s medium (DMEM) supplemented with 10% fetal bovine serum (FBS) and 1% penicillin + streptomycin antibiotics (Gibco, Grand Island, NY) with 5% CO_2_ at 37 °C. To generate stable cell lines, cells were co-transfected with a variant PMP22-myc^exo^ plasmid and modified piggyBac transposase ^39^. Twenty-four hours after transfection, PMP22-myc^exo^ expressing cells were selected in the media containing 5 µg/ml puromycin for 2 days (Sigma, St. Louis, MO). Cells expressing mTAGBFP2 (and PMP22-myc) were sorted from non-mTAGBFP2 expressing cells by fluorescence-activated cell sorting then cultured in the absence of puromycin prior to measuring the cell surface levels of PMP22-myc^exo^.

PMP22 protein expression levels were assessed by immunoblot analysis of cell lysates generated from stable FACS-sorted cell populations. Cell extracts were made by lysing cells 30 min on ice in PBS buffer pH 7.4 supplemented with 1% DDM, 0.2% CHS detergents (Anatrace, Maumee, OH) and cOmplete^TM^, Pefabloc® protease inhibitors (Sigma, St. Louis, MO). After depleting nuclei and cell debris by centrifugation at 12,400 rpm for 15 min at 4 °C, the post-nuclear supernatants were mixed with SDS-sample buffer. Total protein concentration was determined by BCA assay (Pierce). Equal amounts of proteins were separated by SDS-PAGE and immunoblotted using mouse anti-myc-Tag (9B11) antibodies (Cell Signaling Technology, Danvers, MA) and mouse anti-b-actin antibodies (Invitrogen). ImageJ2 software was used to measure density of PMP22-myc protein bands.

To quantify the relative levels of PMP22-myc^exo^ at the cell surface, sorted stably-expressing cell populations were lifted from tissue culture plates with Versene solution (Gibco), pelleted and resuspended in PBS supplemented with 0.5% BSA, 0.1% sodium azide then incubated with mouse anti-myc-Tag (9B11) primary antibody (Cell Signaling Technology, Danvers, MA) for 1 hr on ice, washed at 4°C, and labelled with goat anti-mouse Alexa 568 fluorescent secondary antibody as previously described ^23^. Flow cytometry data were obtained using a Becton Dickinson LSRII equipped with 405 nm, 488 nm, and 561 nm lasers. Analysis and data graphing was done using FlowJo v10 software (FlowJo, Ashland, OR 97520). The gating strategy used FSC and SSC gates to define the median range of cell size and quality. A gate of high BFP expression was used to compare levels of myc/Alexa 568 labeling across cell lines with a similar range of BFP expression as a fiduciary marker for PMP22∼BFP mRNA levels based on flow cytometry analysis of the selected cell population.

## Results

### Demographics

Fifty patients from 35 families were identified with *PMP22* sequence variants (Tables 1 and 2 and Supplementary Tables 1 and 2). Patients with the p.T118M variant were excluded from this cohort because the clinical consequences of this variant remain unresolved ^40^, including one patient who had a p.T118M variant in trans with a *PMP22* deletion. Two additional individuals were excluded from natural history components of this study as their variant classification and clinical presentation were not consistent with CMT1E, although their associated variants (p.Q63R and p.R159H) were included in the variant distribution analysis. The remaining 48 patients had a CMT1E phenotype (Supplementary Table 1) and were included in the CMT1E natural history analysis. The baseline clinical presentations included six patients who presented with hearing loss, 11 with scoliosis, three with hip dysplasia, and one with both scoliosis and hip dysplasia. Except for the patients with p.A106V and p.S131C, the patients with likely pathogenic or pathogenic variants (LP/P) had slowed ulnar motor conduction velocities (Supplementary Table 1).

**Table 1.** Demographic Information and Prominent Clinical Presentation Features. For all PMP22 sequence variants, demographic information was collected at the patient’s baseline evaluation, including their biological sex and age. The Rasch-modified CMT Examination score version 2 (CMTESv2-R) was collected at the baseline and follow-up visits to assess disease progression. The CMTESv2-R scores are separable into <9 (mild), 10-18 (moderate), or >19 (severe). For individuals under the age of 20, the CMT Pediatric Scale (CMTPedS) was performed and can be further separated into <14 (mild), 15-29 (moderate), or >29 (severe). For patients in the CMT1E cohort, additional clinical information was collected at baseline and follow-up visits, including developmental walking milestones, hearing loss, scoliosis, and hip dysplasia.

| Characteristics | Baseline Values | Final Values |
| --- | --- | --- |
| **PMP22 Variants** |  |  |
| **Demographic Information** |  |  |
| Number of variants | 24 |  |
| Number of patients | 50 |  |
| Number of families | 35 |  |
| Females; Males | 29; 21 |  |
| Age range; mean (years) | 3 – 69; 28.43 ± 19.94 |  |
| **CMTEsv2-R** |  |  |
| Standard response mean | 0.49 |  |
| Number of patients | 46 | 29 |
| Age Range; mean (years) | 4-69; 30.52 ± 19.30 | 12 – 79; 39.76 ± 20.24 |
| Score range; mean (out of 32) | 1 – 31; 16 ± 7.72 | 1 – 31; 18.72 ± 7.19 |
| **CMTPedS** |  |  |
| Standard response mean | 0.64 |  |
| Number of patients | 17 | 7 |
| Age Range; mean (years) | 4 – 18; 11.76 ± 5.48 | 13 – 20; 16.86 ± 2.67 |
| Score range; mean (out of 44) | 16 – 37; 28 ± 6.35 | 22 – 38; 33.57 ± 6.63 |
| **CMT1E Cohort** |  |  |
| **Demographic Information** |  |  |
| Number of variants | 22 |  |
| Number of patients | 48 |  |
| Number of families | 33 |  |
| Females; Males | 29; 19 |  |
| Age range; mean (years) | 3 – 69; 27.06 ± 19.29 |  |
| **Clinical Features** |  |  |
| Hearing loss | 6 (12.5%) | 11 (22.92%) |
| Scoliosis | 11 (22.92%) | 13 (27.08%) |
| Hip dysplasia | 3 (6.25%) | 2 (4.17%) |
| Scoliosis and hip dysplasia | 1 (2.08%) | 3 (6.25%) |
| Never walked independently | 5 (10.42%) |  |
| Delayed walking milestones | 29 (60.42%) |  |
| Met walking milestones | 12 (25%) |  |
| Unknown walking milestones | 2 (4.17%) |  |

### *PMP22* Classification, Domain Distribution, and Clinical Phenotype

Among these 48 patients, 24 different *PMP22* variants were identified, including 19 missense variants, three deletions, and two truncations. Twenty-two of these variants were classified as LP/P) (Table 2), and were not found in gnomAD v.3.1.2. Twenty variants have been previously reported ^6, 7, 41–56^. Four variants are novel and have not been previously published, including p.L82P, p.A106V, p.A113P, and p.Q63R. The single amino acid missense variants were graphically mapped onto a 2D schematic map of PMP22 (Fig. 1A) and a 3D structural model (Fig. 1B); 20 of the 24 variants mapped to the transmembrane (TM) domains, 10 of which were within the second TM domain (Fig. 2C). Three variants (p.W28R, p.Q63R, p.S131C) mapped to the extracellular of the protein, including two associated with a mild phenotype (p.W28R and p.S131C) and one (p.Q63R) with an HNPP-like phenotype. The R159H variant, of uncertain pathogenicity (see below), mapped the intracellular space. The remaining five variants were partial gene deletions or truncations (p.Phe84del, p.Cys85*, p.Gly107fsVal*4, p.Ala115_Thr118del, Exon 4 del) (Supplementary Fig. 3), and were not included in the cell surface trafficking and topological correlations studies.

**Figure 1.**
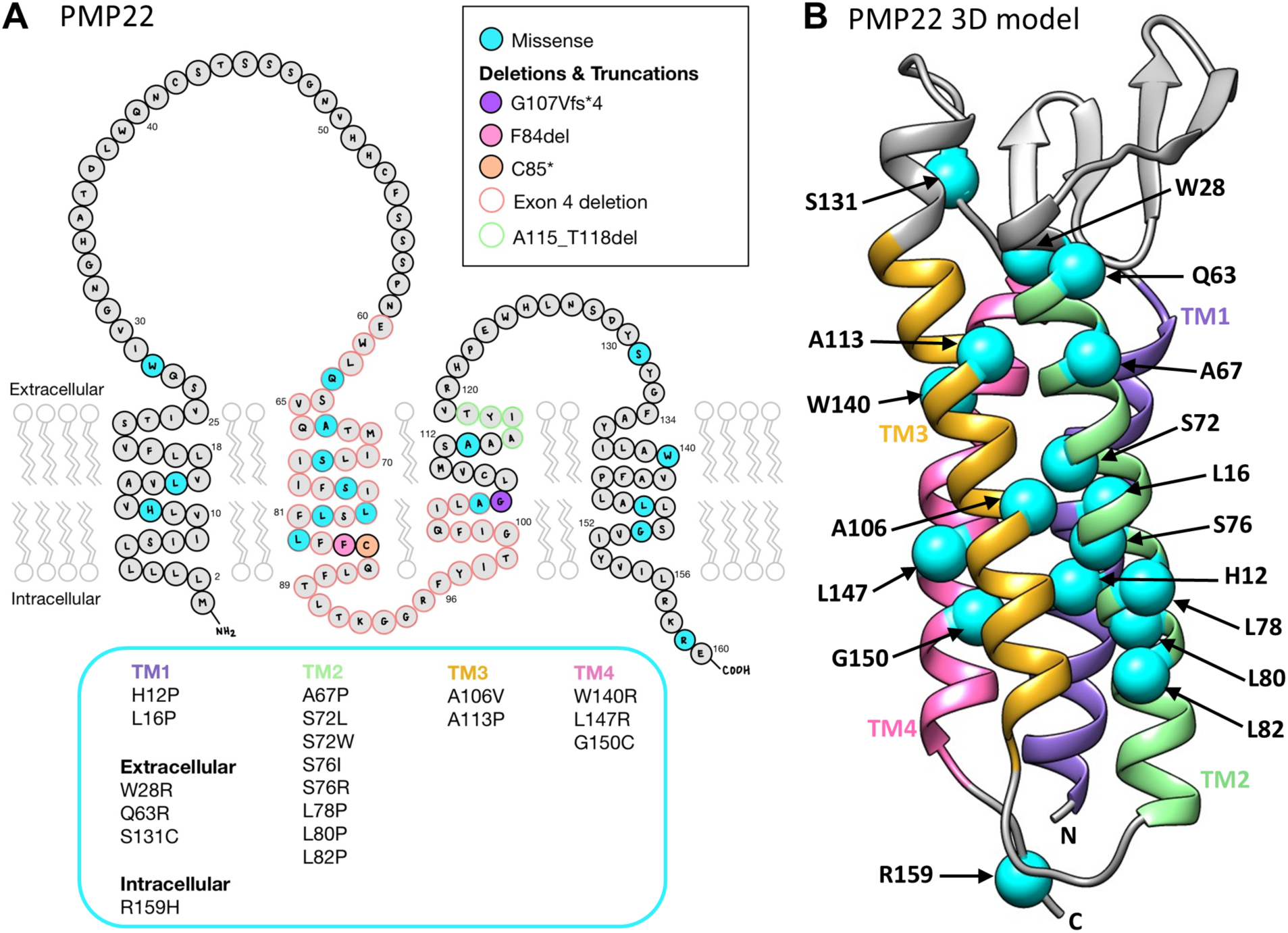
Genotype and structure descriptions of CMT1E patient *PMP22* gene variants included in this study. **(A)** The protein schematic depicts the full sequence and transmembrane topology of human PMP22 with amino acid positions of single point mutations and deletions colored. Patient variants are listed by protein region. **(B)** The predicted 3D folded structure of PMP22 illustrates the four transmembrane helical bundle. Locations of patient variants are denoted by their alpha carbon only (cyan) and labelled.

**Figure 2.**
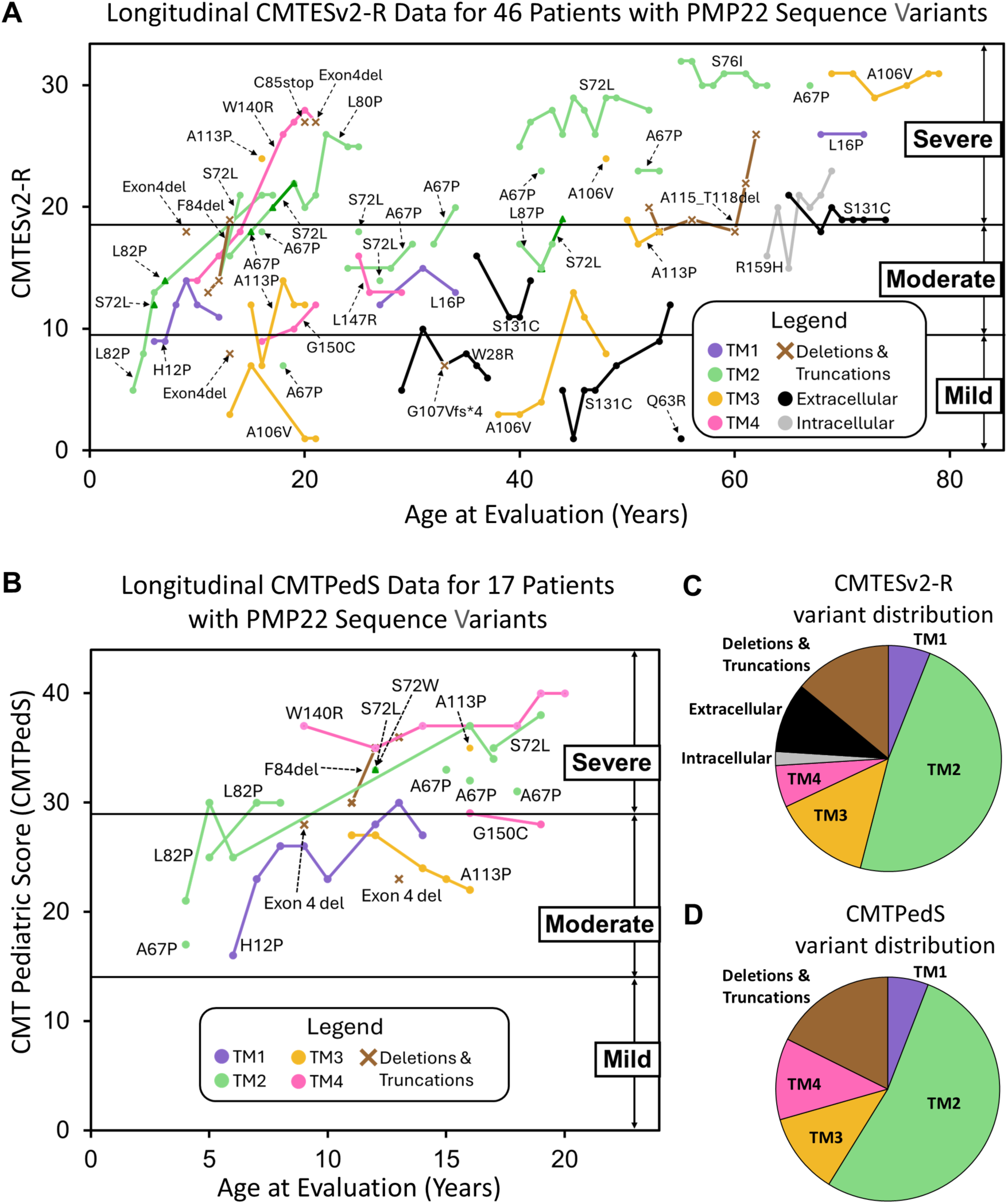
Phenotype evaluation of patients with PMP22 sequence variants using clinical outcome measures. **(A)** Rasch modified CMT exam score version 2 (CMTESv2-R) measuring PMP22 sequence variant disease severity for individual patients are plotted by age, labelled by patient variant, and colored by protein region. **(B)** For patients under age 20, the CMT Pediatric Scale (CMTPedS) was performed to measure the patient’s muscle strength, balance and coordination, and upper and lower extremity function. These scores are plotted by age at evaluation, labeled by patient variant, and colored by protein region. **(C)** The distribution of PMP22 sequence variants across transmembrane helices and extracellular/intracellular regions for patients who completed the CMTESv2-R. **(D)** The distribution of PMP22 sequence variants across transmembrane helices for patients who completed the CMTPedS.

**Table 2:**
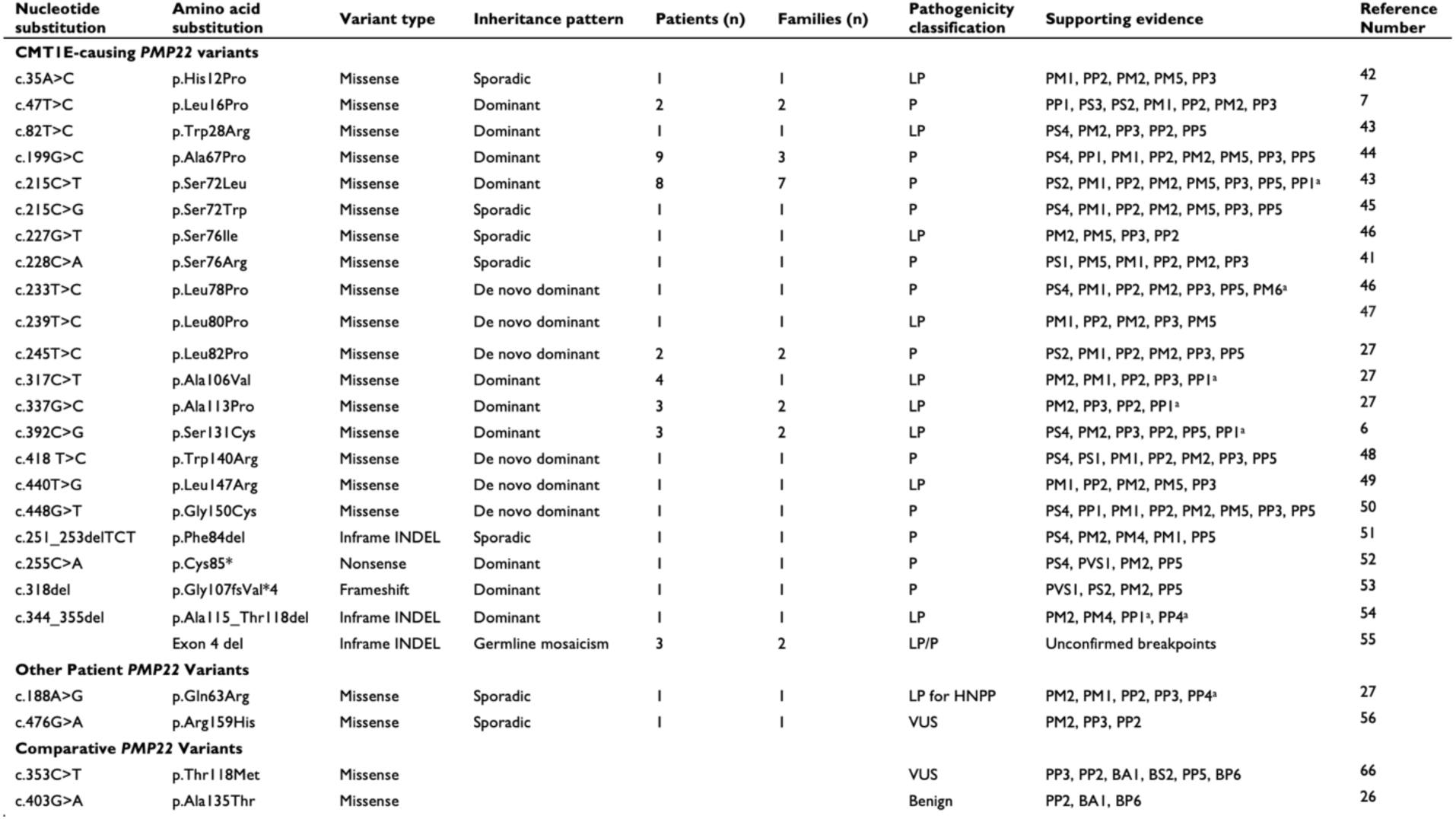
*PMP22* variant classification. ^a^Additional classification criteria added with evidence from this study. (PP1) Pathogenic supporting: Cosegregation with disease in multiple affected family members in a gene definitively known to cause disease. (PM1) Pathogenic Moderate: Non-truncating non-synonymous variant is located in a mutational hot spot and/or critical and well-established functional domain. (PVS1) Pathogenic Very Strong: Null variant in a gene where loss of function is a known mechanism of disease. (PP2) Pathogenic Supporting: Missense variant in a gene with low rate of benign missense mutations and for which missense mutation is a common mechanism of a disease. (PM2) Pathogenic Moderate: Extremely low frequency in gnomAD population databases with frequency threshold for this gene of <0.05%. (PS2) Pathogenic Strong: De novo in a patient with phenotype consistency, no family history and both maternity and paternity are confirmed. (PP3) For a missense or splicing region variant, computational prediction tools unanimously support a deleterious effect on the gene. (PS3) Pathogenic Supporting: Well-established functional studies show damaging effect on the gene or gene product. (PM4) Pathogenic Moderate: Protein length changes resulting from in-frame deletions/insertions in a non-repeat region or a stop-loss variant. (PS4) Pathogenic Strong: For dominant rare disorders, appeared in affected cases while extremely rare in population. (PP5) Pathogenic No Influence: Reputable source recently reports variant as pathogenic, but the evidence is not available to the laboratory to perform an independent evaluation so not included for classification. (PM5) Pathogenic Supporting: Different amino acid change as a known pathogenic variant. (PM6) Pathogenic Moderate: De novo in a patient with phenotype consistency, no family history and both maternity and paternity are assumed. (PS1) Pathogenic strong: Same amino acid change as a previously established pathogenic variant regardless of nucleotide change. (PP4) Pathogenic supporting: Patient’s phenotype or family history is highly specific for a disease with a single genetic etiology. (BA1) Benign Strong: MAF is too high for disorder. (BS2) Benign Strong: Observation in controls inconsistent with disease penetrance. (BP6) Benign Supporting: Reputable source w/out shared data = benign.

The patient with the p.Q63R variant has a clinical presentation that is consistent with Hereditary Neuropathy with Liability to Pressure Palsies (HNPP). Unpublished data highlights multiple families with this variant and phenotype, classifying this variant as LP. Additionally, this variant traffics similarly to wild type (WT) PMP22 in transfected cells (Fig. 4C), further supporting this conclusion.

The patient with the p.R159H variant had symptom onset in the 60s, with absent touch, pinprick, and vibration sensation in the lower extremities up to the ankle, a fine rapid tremor bilaterally, and nerve conduction studies characteristic of an axonal and not a demyelinating neuropathy. Segregation studies could not be accomplished in this family. A comprehensive neuropathies next generation sequencing panel, including the sequence analysis and deletion/duplication testing for 72 genes, was otherwise normal.

### Disease Progression Using the Clinical Outcome Assessments

At their baseline evaluation, 15 patients completed both the CMTESv2-R and the CMTPedS evaluation, 31 completed only the CMTESv2-R, and 2 patients completed only the CMTPedS. Two patients did not complete either clinical outcome measure due to young age. In total, 19 patients were evaluated at baseline only and 31 patients were evaluated longitudionally with the number of follow-up years ranging from 2-13 years. Of these 31 patients, 12 were pediatric patients and 19 were adults. This data is further outlined in Supplemenatry Table 1.

One or more CMTESv2-R evaluations were performed on 46 patients, showing a mean score of 16 ± 7.72 (out of 32) at their baseline visit, which is in the moderate range of disability ^33^. For the 46 patients that completed the CMTESv2-R evaluation, 29 were adults, 10 were pediatric patients over the age of 10, seven were under the age of 10. Twenty-five individuals had longitudinal follow up visits with a SRM of 0.49, which is in the moderate range of responsiveness over two year intervals. To further illustrate the disease severity and progression of the different variants, these data were compiled into a longitudinal model (Fig. 2A), which was helpful because longitudinal data were not available for a large portion of each patient’s life, particularly their childhood. The majority of patients had missense variants in the TM domains and tended to present to the clinic early life with CMTESv2-R scores that ranged from moderate to severe. This data also suggests that progression was typically more rapid in childhood. For 10 patients whose visits were all before the age of 20, the SRM was 1.04, which is in the highly responsive range. In comparison, the SRM for a similar numbers of patients in the age range of 21-40, 41-60 and > 61 years old, the SRM was 0.29-0.34, which is in the low responsive range (Fig. 3B). In contrast, the smaller number of patients with missense variants in extracellular or intracellular domains tended to present to the clinic later in life and had CMTESv2-R scores in the mild (0-9) to moderate (10-18) range ^11^.

**Figure 3.**
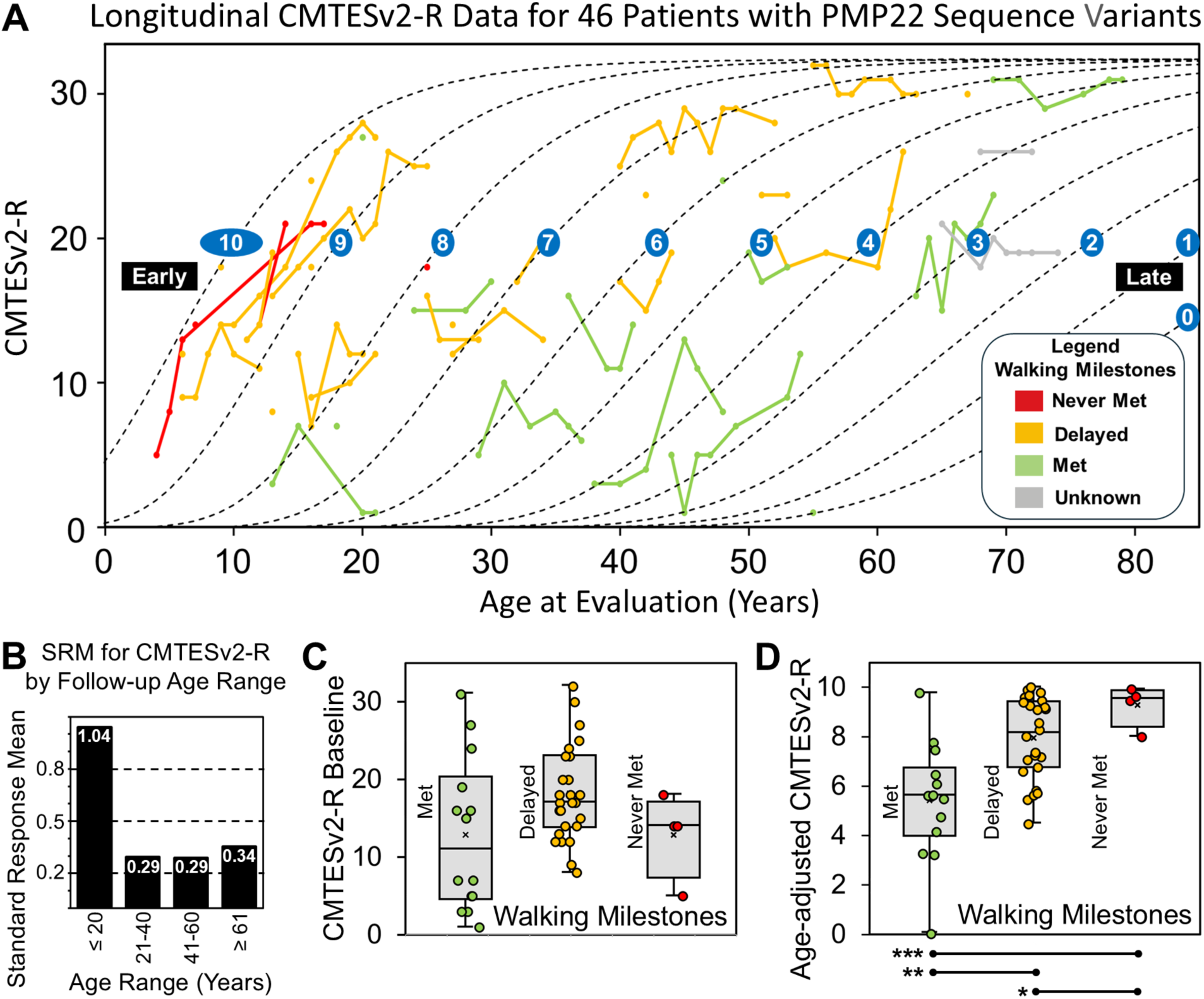
Longitudinal progression of CMT1E patient symptoms. **(A)** Rasch modified CMT exam score version 2 (CMTESv2-R) measuring CMT1E disease severity for individual patients are plotted by age and colored by achievement of walking milestone (variants are labeled in Fig. 2A). Whether a patient met, exhibited a delay in, or never met their walking milestones is inherently tied to the severity of CMT1E symptom progression. Dashed curves follow the age-dependent trajectory of CMTESv2-R progression by a defined equation. Curves are labeled 0 through 10 to indicate their age-adjusted CMTESv2-R value with 0 signifying late CMTESv2-R progression and 10 signifying early CMTESv2-R progression. Fitting of individual patient CMTES-R to the same equation was used to obtain a patient’s age-adjusted CMTESv2-R value (Supplementary Fig. 4). Labels denoting the variant for individual patients can be found in the longitudinal CMT1E patient data plotted in Fig. 2A. **(B)** The standard response mean was calculated for CMTESv2-R and grouped by the patient’s follow-up age. For each CMT1E patient, **(C)** CMTESv2-R baseline values or **(D)** age-adjusted CMTESv2-R values were grouped by walking milestones (Met, Delayed, and Never Met). There is no significant difference in CMTESv2-R baseline values between walking milestone groups (*p*>0.05). In contrast, the age-adjusted CMTESv2-R values are significantly different between walking milestone groups (***=*p*<0.0002; **=*p*<0.002; *=*p*<0.05).

These findings are consistent with the analysis of 17 participants who completed their initial baseline CMTPedS evaluation under the age of 20 (Fig. 2B). A baseline CMTPedS score for these individuals was 28 ± 6.35 (out of 44). A longitudinal analysis on seven of these individuals generated an SRM of 0.64, suggesting moderate responsiveness. All of these patients had moderate to severe neuropathy at their baseline and follow-up visits, reflecting their early disease onset and presentation to the clinic, and all of the missense variants were in TM domains (Fig. 2D). In summary, although fewer patients had missense variants in the intracellular or extracellular domains, they tended to have a later onset and overall milder clinical phenotype than patients with missense variants in the TM domains.

### Genotype-Phenotype Correlation Using Age-Adjusted Measures

The symptoms of patients with *PMP22* variants as assessed by CMTESv2-R generally become more severe over time. The rate of disease progression mirrored whether patients walked independently at or before 15 months, were delayed, or were never able to walk without assistance, as assessed by exam or medical history. Recoloring the longitudinal plot of CMTESv2-R data of individual patients by their walking milestones illustrates the relationship with disease progression (Fig. 3A). Fourteen patients met their developmental walking milestones at or before 15 months of age, 29 patients first walked independently after 15 months, and five patients never walked independently and required walkers or wheelchairs to ambulate. All 29 patients with delayed walking caused by missense variants had variants within TM regions of PMP22, while five of the patients with walking milestone deficiencies had truncations or deletions. Importantly, this measure was consistent across patients who had the same *PMP22* variant (Supplementary Table 1). For instance, all four patients with p.A106V variant met their walking milestone and all eight patients with p.S72L variant had severely delayed walking or never achieved that milestone. These data suggest that the phenotypic consequences of each particular variant are consistent and determine disease progression.

One limitation of the CMTESv2-R score is that it captures a patient’s phenotype only at the time of their clinical evaluation. Without accounting for age, this measure may not accurately reflect disease progression. For example, a patient evaluated in early childhood may have a low CMTESv2-R score despite experiencing early-onset symptoms that could progress to severe disease by adulthood. In contrast, an adult patient with a similarly low score may have a later-onset disease with a milder disease course. This distinction highlights the need to consider age when interpreting CMTESv2-R scores. To extrapolate an age-adjusted measure, the longitudinal CMTESv2-R dataset was modeled to describe changes in disease severity as a patient ages. The sigmoidal trend of disease progression can be approximated by a Gompertz growth curve ^57^ ^58^ (Fig. 3A, Supplementary Fig. 4). The equation of the curve was modified to link a single, age-adjusted CMTESv2-R value to both the midpoint and the slope, which gradually becomes less steep with increasing age. The age-adjusted CMTESv2-R value was spread out so that a value of 10 corresponds to early-onset CMT with rapid progression, and 0 corresponds to a late-onset CMT and slower progression. While the range for age-adjusted CMTESv2-R was set from 0 – 10, roughly matching the full patient range in this study, future CMT1E patients could have values above or below this range.

The validity and utility of incorporating a measure for age-dependent disease-progressionbecame evident when comparing baseline and age-adjusted CMTESv2-R scores to the patient’s developmental walking milestones (Fig. 3C and 3D). Because achieving walking milestones is intrinsically age-dependent, a CMTESv2-R score that captures age-dependent disease progression should be linked to walking milestone measures, which were considered to be met (the ability to walk was achieved by 15 months), delayed (walking achieved after 15 months), and never met.

Baseline CMTESv2-R scores showed no significant differences between met, delayed, and never met walking milestone groups (Fig. 3C). For the four patients who were too young to complete a full CMTESv2-R scores on the initial visit, the first completed CMTESv2-R was used as their baseline score. In contrast, age-adjusted CMTESv2-R values were significantly different for all three walking milestone groups (Fig. 3D).

While CMTESv2-R scores reflect a patient’s symptoms at the time of a visit, they are independent of age. In contrast, age-adjusted CMTESv2-R values allow for normalization across patients, regardless of their age at the time of evaluation. By accounting for age-dependent progression, patients with variants that typically lead to earlier clinical visits can be compared to those whose initial visit occues later in life. Furthermore, using the age-adjusted CMTESv2-R scale enables a more accurate assessment of disease severity across different patients with the same variant. This variant-specific scoring facilitates comparative analysis with the biochemical and cell biological properties of the *PMP22* variant.

The clinical profile of four patients who had a *PMP22* variant that was not a missense variant (Supplementary Fig. 3) was also analyzed. Most of these patients had a phenotype that is more severe than HNPP, which likely results from a toxic gain of function, perhaps the consequences of improper PMP22 folding (see below).

### Cell Surface Trafficking of PMP22 Mutants

Previous studies found that PMP22 variants associated with CMT1E are localized in the ER and not at the cell surface, supporting the idea that misfolded PMP22 is retained in the ER ^7, 8, 10, 59–63^. This analysis was extended to the CMT1E variants in our patient cohort; a benign PMP22 variant (p.A135T) was also used as a comparative negative control. As seen in Fig. 4A, qualitative differences in trafficking to the plasma membrane were found in HEK293 cells expressing GFP-tagged WT PMP22 or the p.S131C variant, which causes a mild phenotype, versus the p.S72L variant, which causes a severe phenotype. WT, p.S131C, and p.S72L variants all colocalized with mTAGBFP2_KDEL, an engineered lumenal marker of the ER, but unlike WT and p.S131C, p.S72L did not appear to be localized to the plasma membrane (Fig. 4A).

**Figure 4.**
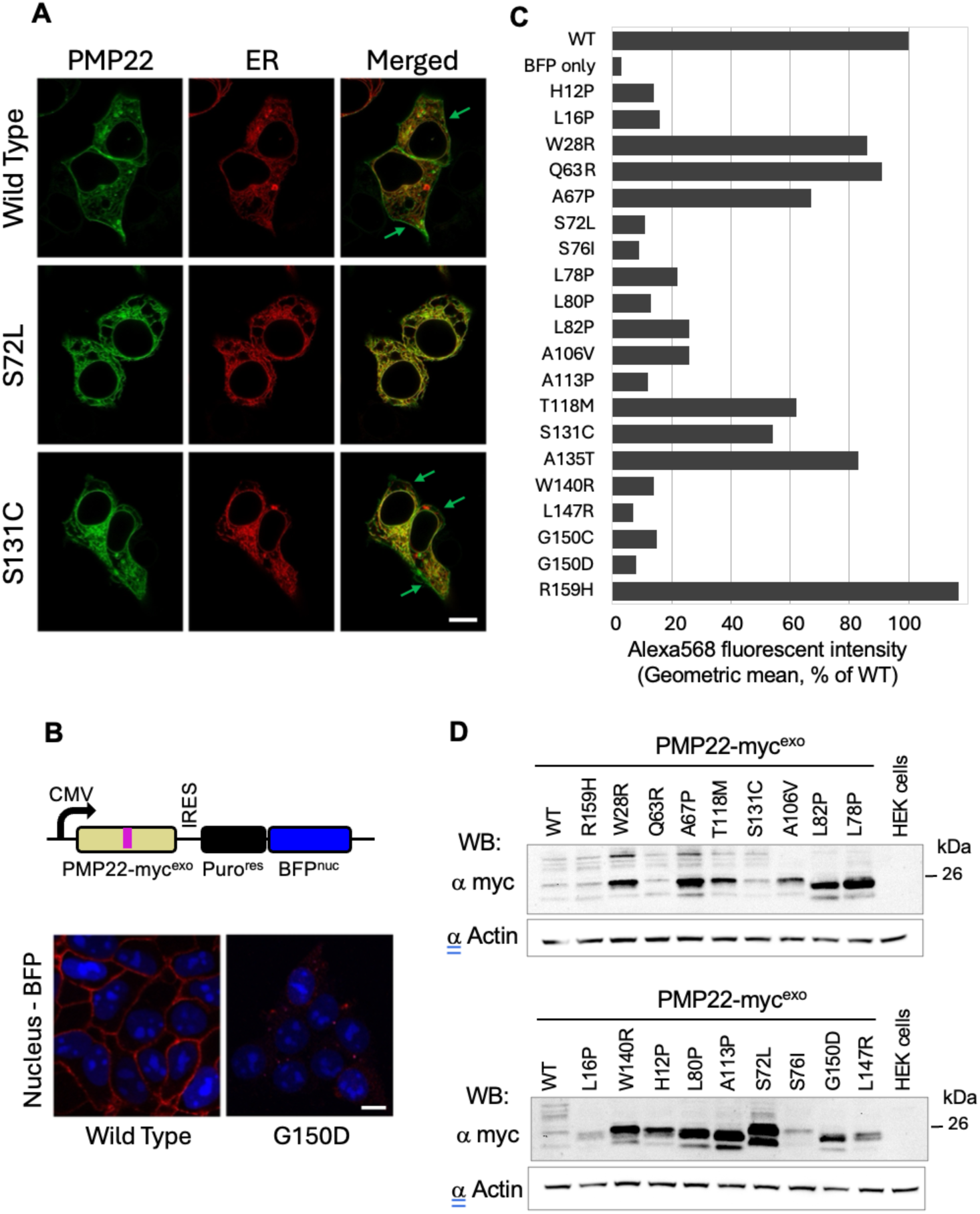
Cell surface trafficking of *PMP22* variants. **(A)** Confocal microscope images of HEK293 cells co-transfected with PMP22-GFP (green) and mTAGBFP-KDEL (depicted in false color red) as marker of Endoplasmic Reticulum (ER). Green arrows highlight PMP22-GFP localized to the plasma membrane. Large black openings in the cells are nucleus. Wild type and S131C PMP22-GFP localized in ER and plasma membrane, while S72L variant PMP22-GFP mostly found in ER. Scale bar: 10 μm. **(B)** PMP22-myc^exo^ expression vector scheme (top panel). The plasmid also expresses a nuclear-localized blue fluorescent protein via an internal ribosome entry site (IRES) within the same mRNA. This allows the expression level of PMP22 to be normalized via the level of BFP. The PMP22 expression plasmids were stably integrated via piggyBac transposase and selection with puromycin. Low panel - confocal microscope image of a stable PMP22-myc^exo^ HEK293 cell line with immunofluorescent cell surface labeling as used for flow cytometry experiments. Wild type PMP22-myc^exo^ can be found localized to the cell surface (in red) but not G150D mutant protein. **(C)** Cell surface localization of PMP22-myc^exo^ variants analyzed after Flow cytometry surface immunostaining of HEK293 stable cell lines. Graph bars represent geometric mean of Alexa 568 fluorescence intensity for tested PMP22 variants normalized to the wild type. **(D)** PMP22-myc^exo^ protein expression in the cell lines used for Flow cytometry experiments was tested by Western blotting with anti-myc antibodies. Equal amounts of cell lysates were loaded onto SDS-PAGE. Anti-b-actin antibody were used as a loading control. Full variant data values are available in Supplementary Table 2.

To assess cell-surface trafficking quantiatively, the cell-surface expression of an epitope-tagged PMP22 (PMP22-myc^exo^) was measured, which carries a c-myc epitope in the second extracellular loop, allowing the PMP22-myc^exo^ to be labeled with α-myc antibodies in non-permeabilized cells ^23^. This assay builds on previous work demonstrating that the extracellular myc epitope does not induce PMP22 aggregation and that PMP22-myc^exo^ surface abundance reflects the known effects of p.L16P and other PMP22 variants that are retained in ER compartments ^19, 22, 64^. The PMP22-myc^exo^ proteins were expressed in tandem with a nuclear-localized blue fluorescent protein (BFP^nuc^) and the puromycin resistance protein (PUR^r^) via an internal ribosome entry site (IRES) (Fig 4B). These expression cassettes were stably integrated into the genome using the piggyBac system, allowing for the isolation and analysis of cell populations with uniform BFP^nuc^ levels, providing consistent PMP22 mRNA expression across these cell lines ^23^. This expression cassette includes flanking insulators, providing durable expression through multiple passages of cells ^37^. As a quantitative comparison of the effect of PMP22 variants on cell surface trafficking, several missense variants from our patient cohort were examined by measuring their surface abundance using fluorescent antibodies and flow cytometry. The level of anti-myc / Alexa 568 surface labeling within a subpopulation of cells gated for the same level of BFP^nuc^ was analyzed to ensure similar levels of PMP22_(IRES)Puro^r^_BFP^nuc^ mRNA across cell lines expressing different PMP22-myc^exo^ variants.

Most PMP22 variants showed less cell surface localization than did WT PMP22 (Fig. 4C, Supplementary Fig. 5). Consistent with previous studies, the Tr and Tr-J variants (p.G150D and p.L16P, respectively) as well as other PMP22 variants that are known to be retained intracellularly (p.S72L, p.S76I) ^7, 19, 22, 65^, had very low levels of cell-surface PMP22-myc^exo^ labeling. The p.T118M variant, which was previously observed to have intermediate defects in cell surface trafficking, showed an intermediate defect in cell surface abundance ^66^. Over the whole set of *PMP22* variants, cell surface abundance was found to be variable with some variants showing highly reduced cell surface localization, whereas others were near that of WT PMP22-myc^exo^. In general, the severity of disease correlated with the severity of defect in cell surface localization (see below), consistent with hypothesis that PMP22 intracellular retention is a strong correlate and potential contributor to disease.

In addition, the total cellular steady-state levels of the PMP22-myc^exo^ variants was measured by immunoblotting, as seen in Figure 5. Because each cell population expresses similar levels of BFP^nuc^ (which serves as the fiduciary marker for PMP22 mRNA), differences in the steady-state levels of the PMP22-myc^exo^ variants reflect differences in their rates of synthesis and/or degradation. Fig. 4D and Supplementary Table 2 show a wide range in PMP22-myc^exo^ expression levels across the different variants, likely reflecting differences in the degradation of these variants. There was no apparent correlation between the degree of cell surface trafficking to the overall steady-state level of PMP22-myc^exo^ variants, suggesting that ER retention and ER-associated degradation could be uncoupled. Moreover, the severity of CMT disease phenotypes did not correlate with the steady-state levels of the PMP22-myc^exo^ variants (Supplementary Table 2).

**Figure 5:**
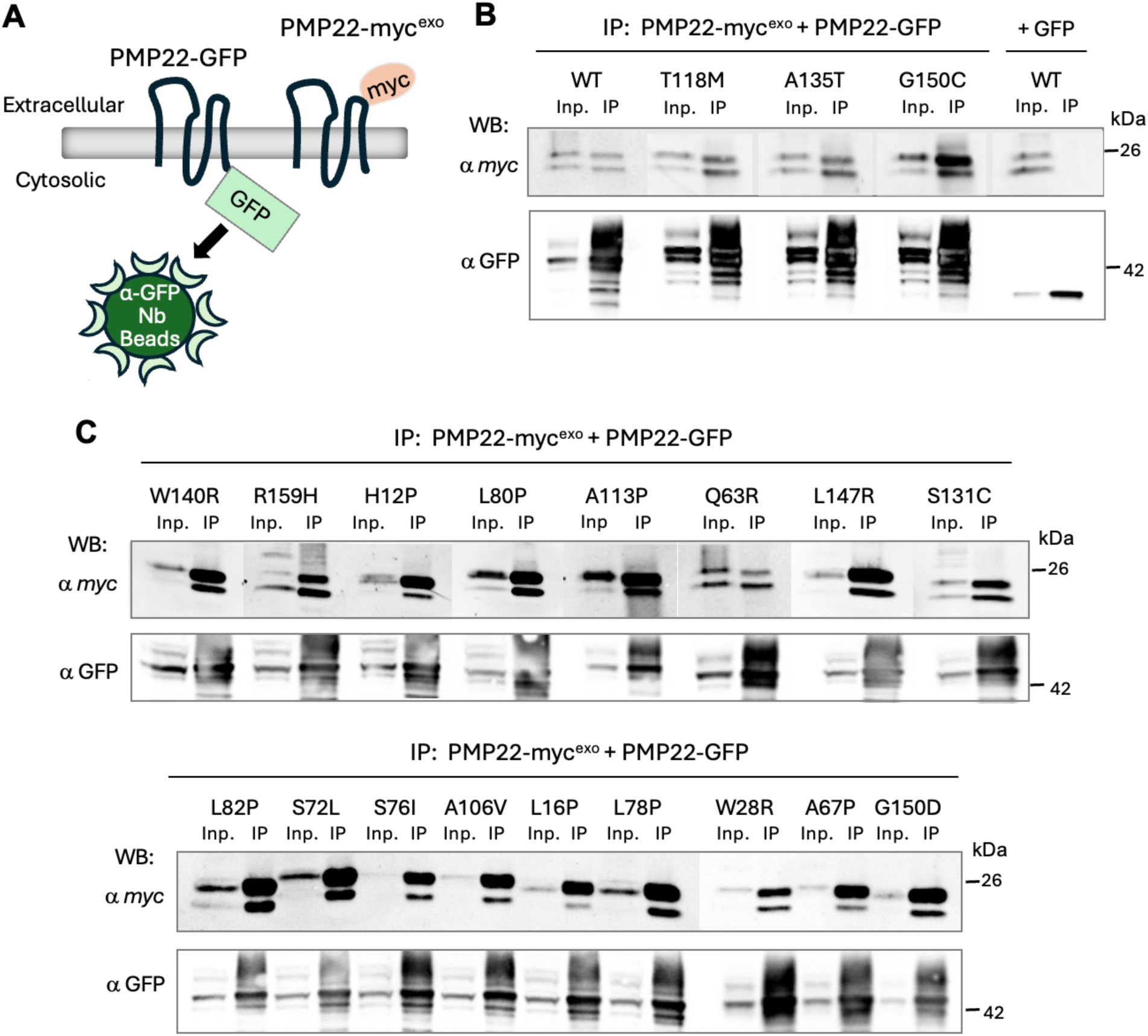
PMP22-myc^exo^ variants can form a complex with PMP22-GFP. **(A)** HEK293 stable cell lines with PMP22-myc^exo^ variants were transfected with wild type PMP22-GFP. After cell lysis, the proteins were extracted and co-immunoprecipitated using nanobody GFP beads (depicted by the cartoon). Eluted with SDS-sample buffer proteins were immunoblotted with anti-myc antibody to detect bound PMP22-myc^exo^ and with anti-GFP antibody to monitor recovered PMP22-GFP. A 5% equivalent of the input lysate was included in the immunoblots. **(B)** Immunoblotting of WT, unsolved *PMP22* variant (p.T118M), negative control variant (p.A135T), and a patient variant (p.G150C). **(C)** Immunoblotting of *PMP22* sequence variants in this cohort and *TrJ* variant (p.G150D).

PMP22 can form dimers, and some CMT1E-causing variants cause higher order oligomers/aggregates ^19–22, 65^. The retention of WT PMP22 in the ER by virtue of its association with mutant ER-retained PMP22 has been hypothesized to contribute to disease pathology ^7, 8, 65,67^. This was investigated by whether the PMP22 variants in our study differed in their ability to oligomerize with WT PMP22 by co-immunoprecipitation from DDM/CHS detergent cell lysates α-GFP nanobody beads ^23^. Figure 5 shows that all PMP22 variants were able to associate with WT PMP22 to similar extent, regardless of their limited cell surface distribution or the disease phenotype correlate. These associations were observed for PMP22 variants that had either strong or limited defects in cell surface trafficking and either caused more or less severe CMT1E.

### Topological Correlations of PMP22 Variants

To determine whether the disease severity associated with different variants was related to their positions within the PMP22 structure, the age-adjusted CMTESv2-R scores were mapped (Fig. 3, Supplementary Fig. 4, Supplementary Table 1) for missense variants onto a structural model of PMP22 AlphaFold model (AF-Q01453-F1-v4) ^28^. As shown in Fig. 6A, variants associated with severe, earlier-onset disease are clustered within the TM regions, particularly in positions that are buried deep within the lipid bilayer (Fig. 6A,C). Linear correlation plots reveal a good correlation between the age-adjusted CMTESv2-R and membrane depth (Fig. 6E). These positions also correlated with reduced cell-surface localization, either as a raw measure (Supplementary Table 2) or normalized to the cell surface expression of PMP22-myc^exo^ (Fig. 6B). Moreover, the positions of variants causing reduced cell-surface localization correlated with the age-adjusted CMTESv2-R (Fig. 6D) and their depth in the lipid bilyer (Fig. 6F).

**Figure 6.**
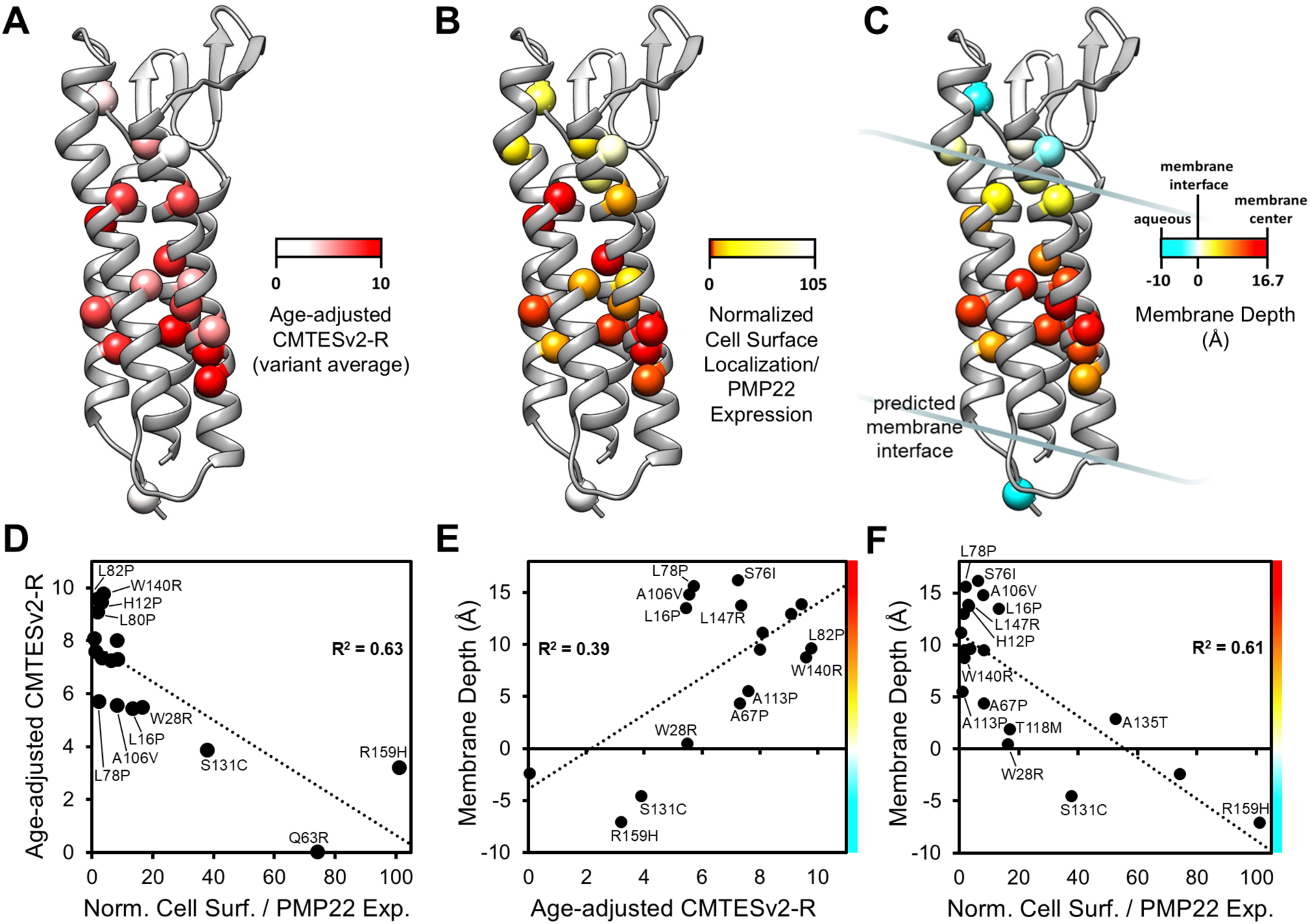
Correlations between CMT1E symptom progression, PMP22 trafficking, and variant position relative to the membrane. **(A)** Age-adjusted CMTESv2-R values, **(B)** relative expression-adjusted PMP22 cell surface localization, and **(C)** predicted depth in the membrane bilayer are mapped onto the PMP22 structure to depict the visual correlation between these variant parameters in 3-dimensional space (variant positions are labeled in Fig. 1B). The membrane depth of each variant position was derived from the membrane-embedded position of the PMP22 AlphaFold model as predicted by the PPM method. For variants, plots of the linear correlation for **(D)** CMT1E symptom progression and PMP22 cell surface localization, **(E)** CMT1E symptom progression and predicted depth in the membrane bilayer, and **(F)** PMP22 cell surface localization and predicted depth in the membrane bilayer. For each correlation plot, R^2^ are indicated for the linear fits (*p*<0.01 for each) and only variants with |standardized residuals| > 0.5 are labeled. Full variant data values are available in Supplementary Table 2.

These structural correlations were compared to how variant positions would be predicted to cause disease using AlphaMissense (https://alphafold.ebi.ac.uk/entry/Q01453) ^68^, a deep-learning model that predicts the pathogenicity of missense variants, and found that substitutions at positions within the TM regions were “likely pathogenic” (LP). These results underscore the importance of the TM regions in disease progression and further demonstrate how the disruption of both the TM helix packing and interaction with their surrounding lipidic environment are important for normal cell surface trafficking.

## Discussion

CMT1E has been characterized by a wide variety of clinical phenotypes related to different *PMP22* variants. A few *PMP22* missense variants reported as CMT1E are associated with cases that resemble HNPP and are likely cause loss of function (as exemplified by the p.Q63R variant reported here), and other missense variants cause a CMT1A-like phenotype ^69^. Most of the 46 CMT1E patients reported here presented with significant neuropathy in infancy, and could also be classified as Dejerine-Sottas disease ^5, 70^. The onset of independent walking was delayed in 29/46 patients, five were never able to walk independently, and only 16 met their normal developmental milestones. Of the 17 subjects who were evaluated in childhood by the CMTPedS scale, all but one progressed into the severe range of impairment by age 15. As demonstrated by SRM, progression in many patients was more rapid prior to the age of 20 (SRM 1.04) compared to adulthood (SRM 0.29-0.34 from ages 21 to >60).The same variants caused similar impairment in different patients, supporting prior studies that the type and nature of specific *PMP22* variants determines the severity of CMT1E ^5^.

Longitudinal data showed progressive disease for all patients, either by the CMTESv2-R or by the CMTPedS. As an additional measure of progression, the CMTESv2-R score as a function of age was modeled to extrapolate an age-adjusted CMTES score (Fig. 3). The age-adjusted CMTESv2-R values broadly correlated with the walking milestone score. Patient groups with normal or delayed walking milestones had an average age-adjusted CMTESv2-R values of 5.4 and 7.9 out of 10, respectively, whereas the average value for patients that never achieved their walking milestone was 9.3. This study found that walking milestones are a useful marker for disease severity during a specific developmental age range. The onset of disease severity for patients that shared the same *PMP22* variant was similar by both the age-adjusted CMTES values and the walking milestone score, reinforcing the view that the severity of CMT1E at a given age is largely determined by the specific *PMP22* variant. While the current model of disease progression is limited to less than 10 years, future studies will aim to follow the long-term progression of patient symptoms to improve the model’s accuracy as a predictive tool for individuals with genetic diagnoses.

All 20 variants within the TM domains caused moderate to severe CMT. This study shows for the first time that the localization of the variant within the PMP22 TM domains predicts disease severity. Moreover, the predicted depth of the variant positions within the lipid bilayer, as well as the hydrophobic density of the lipidic environment surrounding variants, correlated with progression of disease severity. Ten of these variants were in TM2, more than twice as many as were found in other TM helices domains. As previously demonstrated, the TM2 region mediates the interaction between PMP22 and MPZ, the major adhesion protein in compact myelin ^23^. Disrupting this interaction might contribute to the pathogenesis of these neuropathies.

Four of the 24 missense variants in our cohort mapped outside the TM domains (Fig. 1A). The p.R159H variant is unlikely to cause disease even though it was unanimously predicted to have a deleterious effect by computational prediction tools (PP2). The phenotype of the affected patient does not fit within the known phenotypes associated with *PMP22*-related neuropathies.

Furthuremore, the p.R159H variant traffics to the cell surface at least as well as WT PMP22, and the variant is found too frequently in gnomAD v.3.1.2 (allele count 22; 1.36x*e*^−5^) to the cause of a disease as rare as CMT1E. A family with a different amino acid substitution at the same codon (p.R159C) was reported to have an axonal neuropathy ^56^, but this variant is also found at a high frequency in gnomAD v.3.1.2 (allele count 22; 1.36x*e*^−5^). In contrast, none of the CMT1E variants reported here were found in gnomAD v.3.1.2.

The three missense variants (p.W28R, p.Q63R, and p.S131C) within the extracellular domain cause neuropathy. Both p.W28R and p.S131C are associated with milder, CMT1-like phenotypes than the variants located in TM domains. Whether the dominant effect of these variants is similar to that of the TM domain variants remains to be determined. Because the p.Q63R variant causes an HNPP-like phenotype (a much milder phenotype than even typical CMT1A), it likely causes a loss of function rather than a toxic gain of function.

Myelinating Schwann cells express large amount of PMP22 protein. Most of newly synthesized PMP22 is retained in the ER and degraded with only a small proportion reaching compact myelin^15^. Variant forms of PMP22, including *Tr* (p.G150D) and *TrJ* (p.L16P) are retained in the ER both in vitro and in vivo ^7–10, 19, 21, 22, 61, 62, 64^. These variants can also complex with WT PMP22, sequestering it from the plasma membrane and accumulating it in the ER ^8, 20^. Such a sequestration model has been suggested as a potential disease mechanism ^67^. This study confirms and expands upon these findings with the missense variants found in our patient cohort, by showing that retention in the ER, sequestration from the cell surface, and the ability to interact with wildtype PMP22 are hallmarks of variants that cause severe disease. Although intracellular retention has been suggested to be a pathogenic mechanism of CMT1A and CMT1E ^7, 19, 62, 71^, how retented PMP22 causes cellular defects remains unclear. Possibilites include sequestering ER chaparones ^72, 73^, sequestering cholesterol ^64, 74–78^, inducing maladaptive features of the unfolded protein response (UPR), as described for retained *MPZ* variants that cause CMT1B ^79–83^, or complications of PMP22 degradation ^62, 73, 84–88^.

All the PMP22 variants retained their ability to associate with WT PMP22, so that even misfolding, ER retention, and aggregation may not prevent PMP22 mutants from associating with WT PMP22. This is consistent with previous studies showing that *Tr* (p.G150D) and *Tr-J* (p.L16P) PMP22 variants retained their ability to complex with WT PMP22 and potentially sequester it from the plasma membrane ^8, 20^; such a dominant-negative effect has been suggested to be a disease mechanism ^67^. The structural basis for how PMP22 oligomerizes is not known. Structure and function models of a structural relative of PMP22, claudin-15, suggest oligomerization through multiple interfaces, which may explain why no single PMP22 variant tested has abolished homoligomerization ^89, 90^.

This study found that the steady state levels of different CMT1E mutants were variable, which is likely due to differences their degradation rates, as each CMT1E PMP22 variant-expressing cell line was sorted for tandemly-expressed BFP to ensure comparable expression levels. It was also surprising that some variants with high steady state levels did not correlate with their disease severity. The disease mechanisms mentioned above should, in theory, worsen with higher levels of the PMP22 variants. One caveat is that HEK293 cells may have a different cohort of degradative machinery and pathways than do myelinating Schwann cells, which have been hypothesized to use ER-associated degradation and lysosomal sorting/autophagy to degrade PMP22 ^15, 61, 72, 91–94^.

In summary, this analysis found that most cases of CMT1E are caused by pathogenic *PMP22* variants located in TM domains. Disease severity and rapid progression correlate with the variant’s depth and hydrophobic environment within the respective TM domain, as well as impaired intracellular trafficking from the ER to the cell surface. Studies to further investigate these findings are underway.

## Data Availability

The data that supports the findings of this study are available from the corresponding author, upon reasonable request. The data are not publicly available since they contain information that could compromise the privacy of research participants.

## Acknowledgments

This study was made possible by the personnel at the University of Iowa and the equipment available in the Flow Cytometry facility. Additionally, we would like to express our heartfelt gratitude to the patients and families who participated in this study. Their willingness to contribute their time and experiences made this research possible. We deeply appreciate their trust and support, without which this work would not be feasible. Their involvement has provided invaluable insights that have greatly enriched our understanding of this subject matter.

## Supplementary material

Supplementary material is included here

## AAppendix

We would like to thank study team members for their invaluable contributions to the Inherited Neuropathies Consortium, including Richard A. Lewis, Diana Bharucha-Goebel, Carsten Bonnemann, Francesco Muntoni, Rebecca Traub, Brett McCray, Mario Saporta, Seth Perlman, Vera Fridman, Gyula Acsadi, Hernan Gonorazki, Jun Li, Sabrina Yum, Tim Estilow, Katie Eichinger, Eleonora Cavalca, Luca Crivellari, Franco Taroni, Paola Lanteri, Amedeo De Grado, Daniele Cazzato, Gita Ramdharry, Alex Rossor, Andrea Cortese, Tara Jones, and John Svaren.

---

**Supplementary Figure 1.**
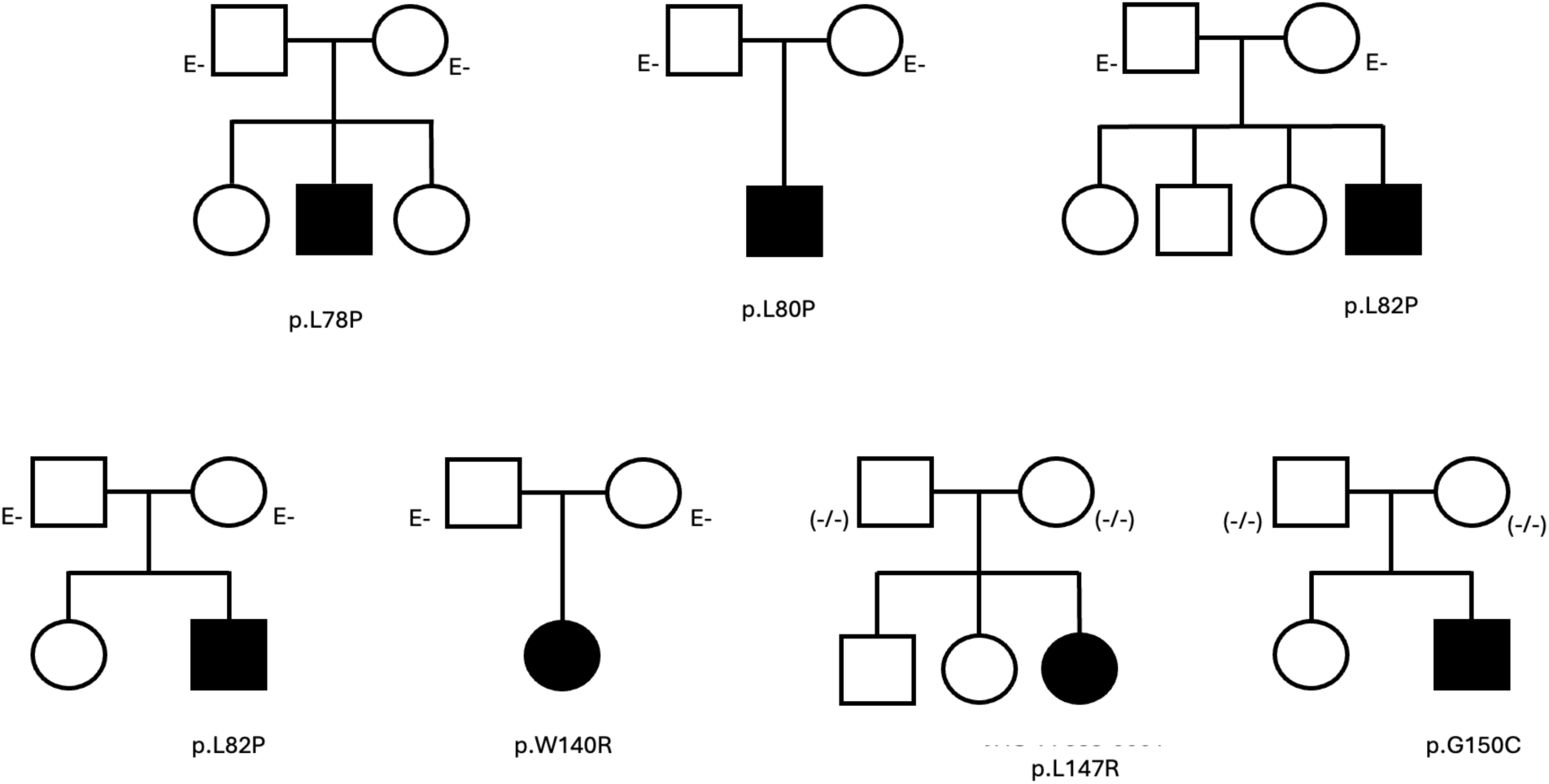
Pedigrees for de novo *PMP22* variants. Squares indicate biological males and circles indicate biological females. Affected individuals are indicated by a shaded in square or circle. Individuals included in this study are indicated by their participant ID and *PMP22* variant. E-indicates the individual was clinically unaffected. (−/−) indicates the individual has negative clinical genetic testing for variant.

**Supplementary Figure 2.**
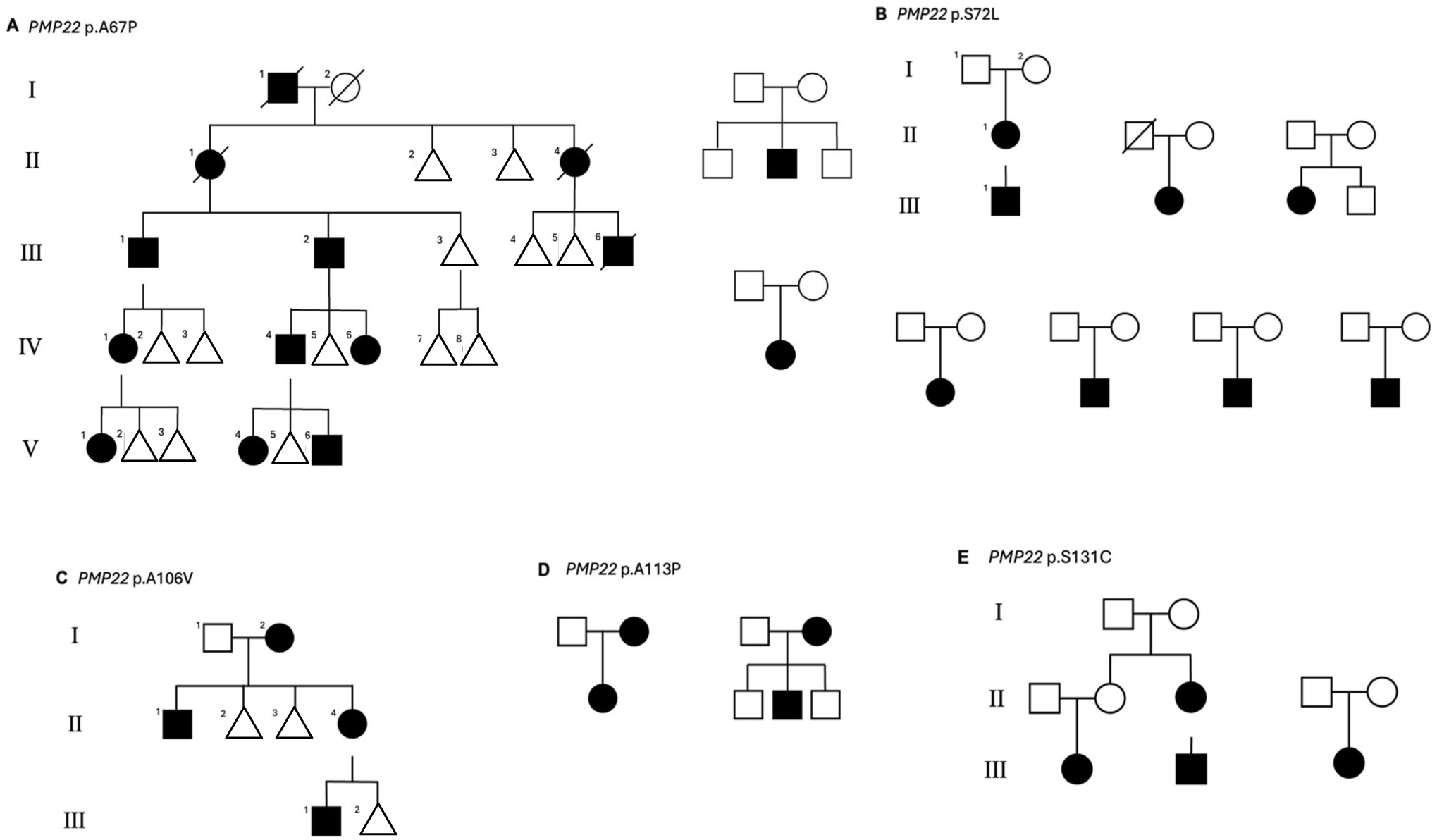
Pedigrees for common *PMP22* variants in our cohort. Squares indicate biological males and circles indicate biological females. Affected individuals are indicated by a shaded in square or circle. For larger pedigrees, the sex of non-affected individuals is masked by using a triangle. Roman numerals (I-V) indicate generation. The number in the top left corner of the shape indicates individual within that generation. A diagonal line through the shape signifies the individual is deceased. Pedigrees are shown for families with the following *PMP22* variants: **(A)** p.A67P, **(B)** p.S72L, **(C)** p.A106V, **(D)** p.A113P, and **(E)** p.S131C.

**Supplementary Figure 3.**
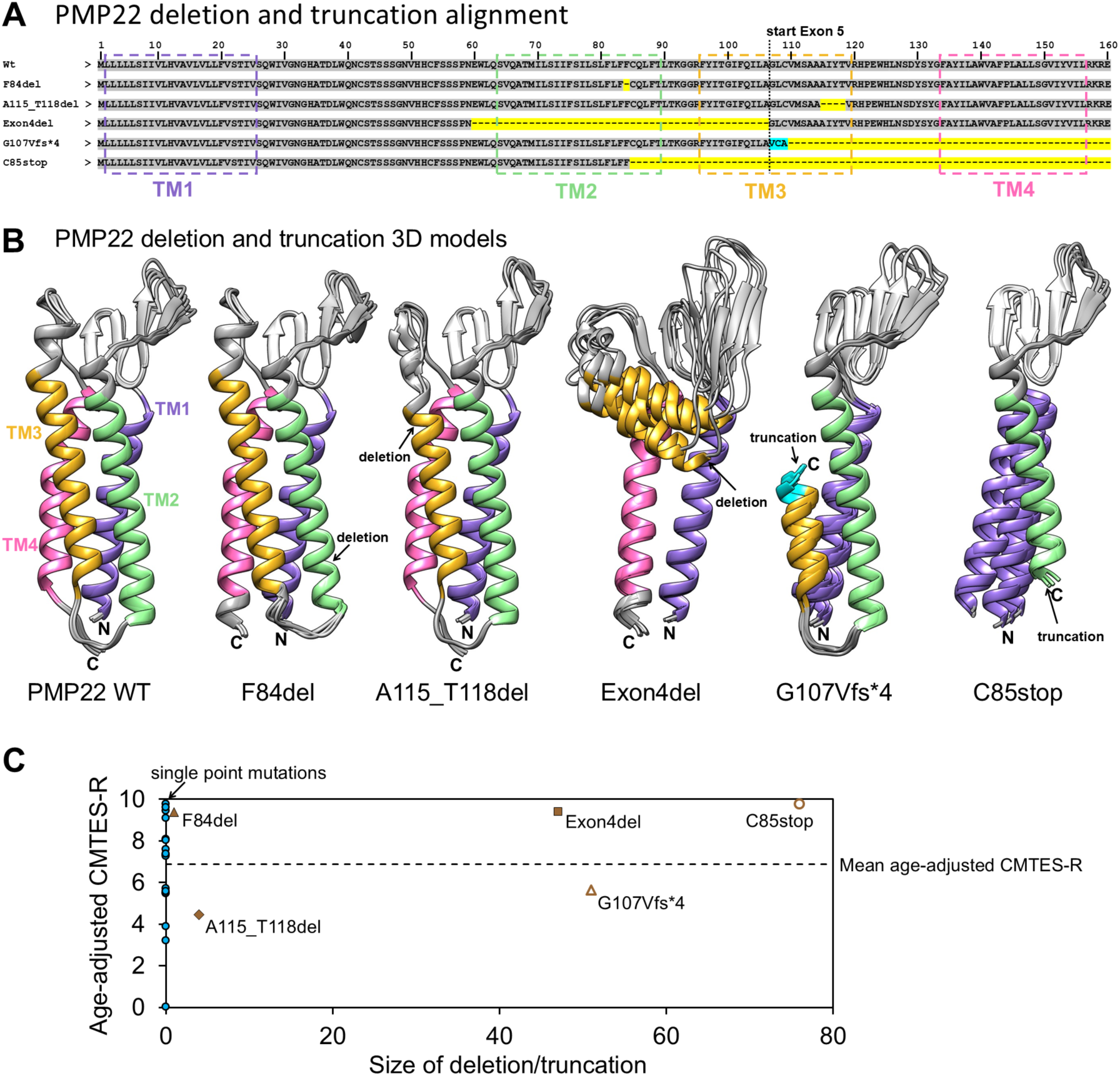
PMP22 deletion and truncation analysis. **(A)** Protein sequence alignment of PMP22 WT, deletions, and truncations for CMT1E patients examined in this study. The expressed PMP22 protein sequences of deletions and truncations were inferred from both exons encoded by WT cDNA and the PMP22 gene sequences of variants from CMT1E patients. Yellow Highlighted regions indicate gaps corresponding to missing amino acids (yellow) or amino acid changes due to a frame shift (cyan). **(B)** Five structural models, generated using AlphaFold3, are aligned for each PMP22 protein sequence and the position of the deletion or truncation is indicated. F84del and A115_T118del retain the basic 4 transmembrane helix bundle, while Exon4del, G107Vfs*4 and C85stop are missing transmembrane helices. These models are only predictions based on expected protein sequence and may not represent the actual cDNA. **(C)** The age-adjusted CMTESv2-R values were determined by fitting longitudinal patient CMTESv2-R values and plotted by the number of missing amino acids in each deletion or truncation. Single point mutations (blue) are included at 0 missing amino acids. PMP22 variants missing large regions are expected to have folding issues and age-adjusted CMTESv2-R values corresponding to early progression of CMTESv2-R symptoms. Interestingly, G107Vfs*4 shows later progression of CMTESv2-R than expected based on the prediction for the expressed protein. Future efforts could address whether the G107Vfs*4 expresses the predicted protein and if it is trafficked to the cell surface.

**Supplementary Figure 4.**
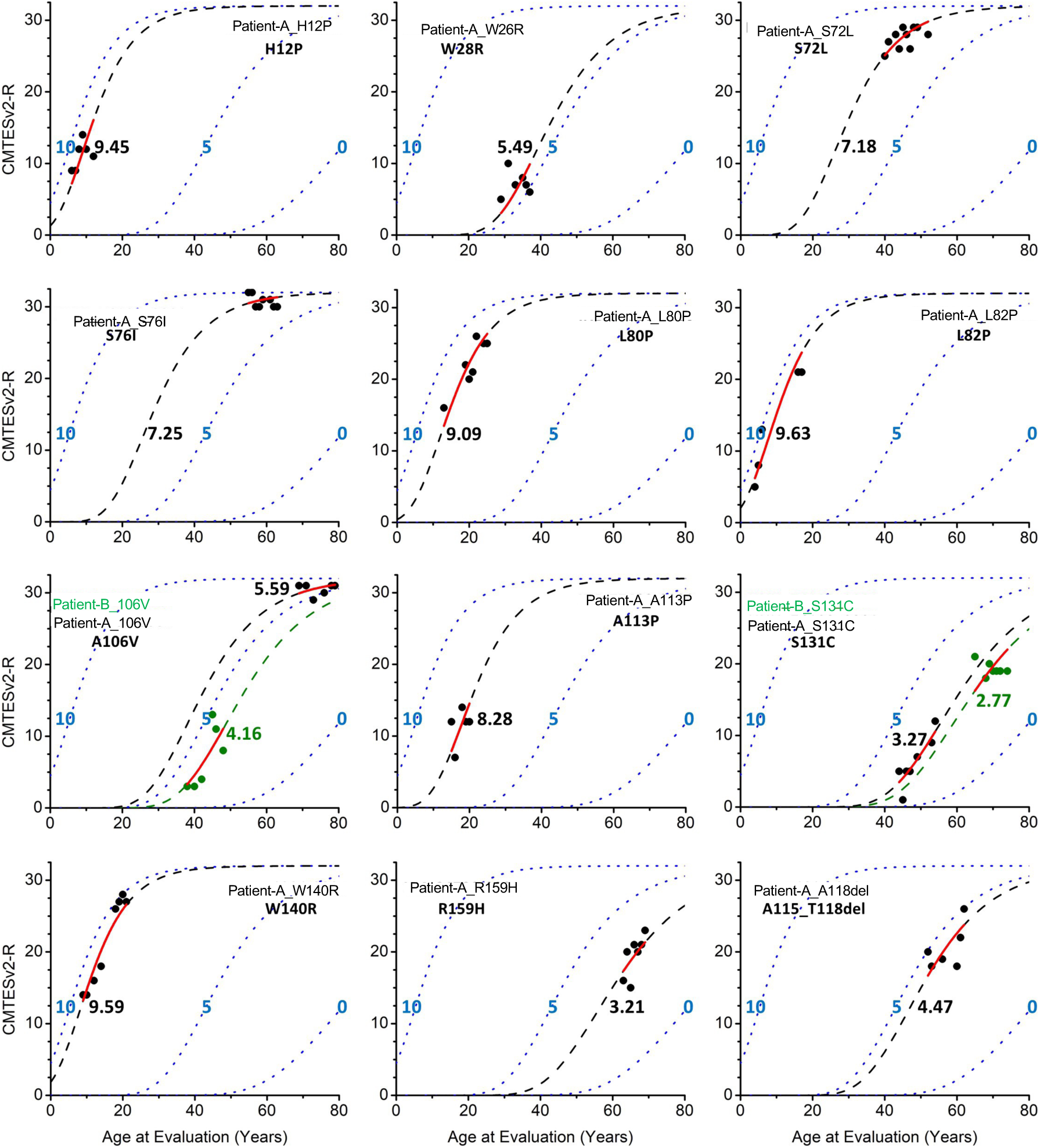
Longitudinal progression and age-adjusted curve fitting for individual patients. For all patients with five or more clinical visits and full assessments, CMTESv2-R scores are plotted as a function of age of evaluation. The CMTESv2-R scores were fit to a modified Gompertz curve and a single fitted value for age-adjusted CMTESv2-R was extracted for individual patients. The fit region is colored in red while the full simulated curve is shown as a dashed line where the age-adjusted CMTESv2-R value is noted next to the curve. For easier comparison of patients with early and late onset of disease severity, simulated, blue-dotted guidelines were set with age-adjusted CMTESv2-R values fixed at 10, 5, and 0. Each plot is labelled with patient local ID and PMP22 variants. For A106V and S131C, two patients are included in the same plot and individual patient CMTESv2-R data points are differentiated by color. For these variants, the age-adjusted CMTESv2-R values are similar despite clinical data representing different age and absolute severity ranges.

**Supplementary Figure 5.**
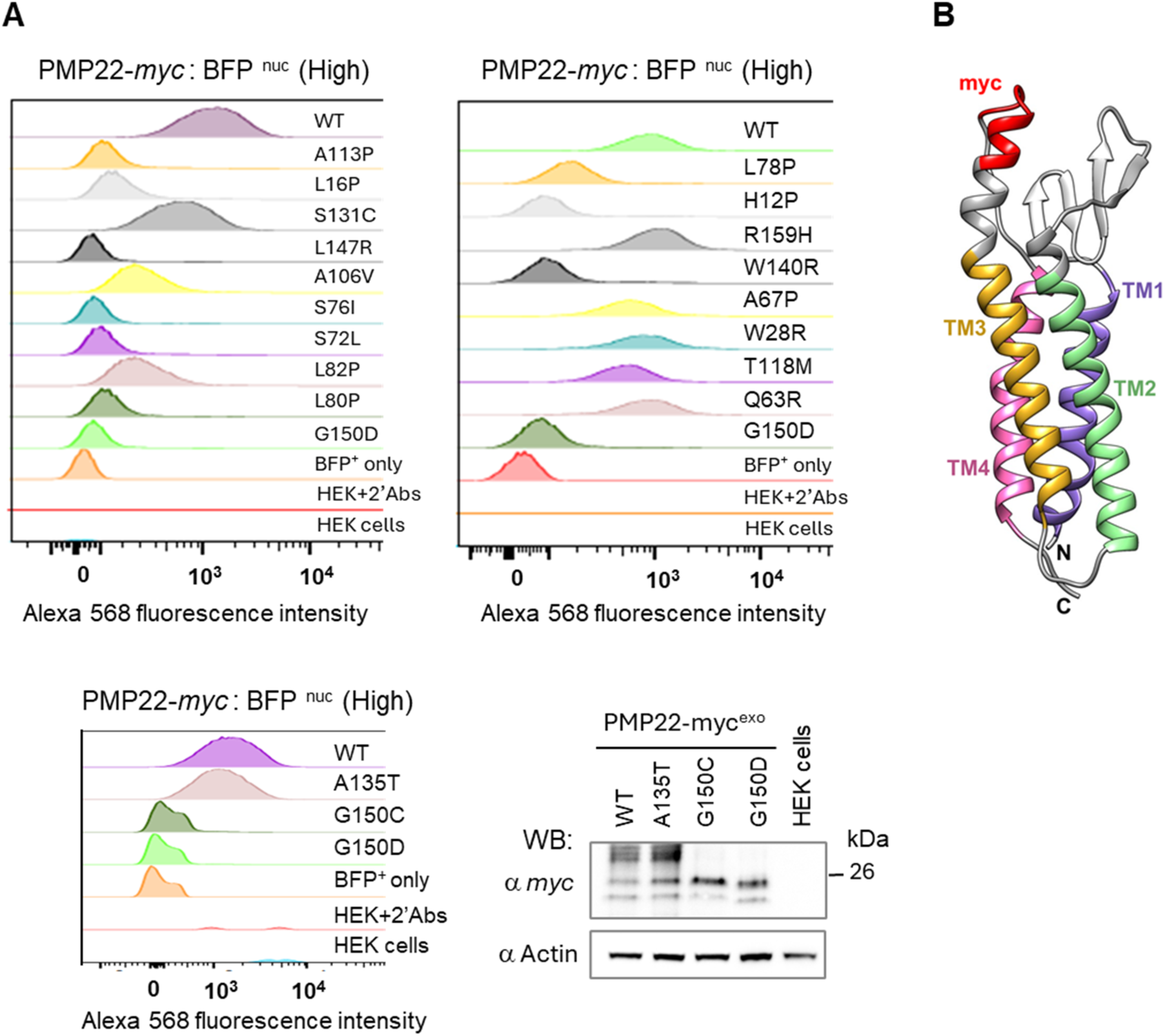
Analysis of PMP22 cell surface trafficking by flow cytometry. **(A)** HEK293 stable cell lines expressing wild type or mutant PMP22-myc^exo^ were labeled with mouse anti-*myc* and goat anti-mouse Alexa568 secondary antibodies. Overlay plots show levels of PMP22-myc^exo^ protein on the cell surface for high expressing PMP22 cell population gated by BFP expression which correspond to the total PMP22 expression in the cells. Expression level across cell lines were normalized by gating for cells with high levels of BFP and then quantified for the level of *myc* epitope exposed on the cell surface. **(B)** PMP22-myc^exo^ structural model with mapped *myc* tag.

**Supplementary Table 2.** Biochemical and cellular analysis of *PMP22* variants correlated with disease severity. *G150C values were used for mapped data onto Figure 6A-C structure.

| **PMP22 variant** | **Topology** | **Average age-adjusted CMTEsv2-R** | **Cell Surface Localization (Normalized to WT)** | **PMP22-myc Aver. Total ProteinWB** | **Cell Surf. Localization / PMP22-myc Aver. Total ProteinWB** | **Membrane depth** | **Protein Surface accessibility** |
| --- | --- | --- | --- | --- | --- | --- | --- |
| H12P | TM1 | 9.45 | 14 | 4.63 | 3.03 | 13.85 | No |
| L16P | TM1 | 5.45 | 16 | 1.21 | 13.28 | 13.47 | Yes |
| W28R | Extracellular | 5.49 | 86 | 5.23 | 16.43 | 0.45 | No |
| Q63R | Extracellular | 0.03 | 91 | 1.23 | 74.20 | -2.42 | Yes |
| A67P | TM2 | 7.32 | 67 | 8.01 | 8.36 | 4.36 | Yes |
| S72L | TM2 | 8.09 | 11 | 17.09 | 0.64 | 11.14 | No |
| S76I | TM2 | 7.25 | 9 | 1.43 | 6.28 | 16.16 | No |
| L78P | TM2 | 5.71 | 22 | 10.11 | 2.18 | 15.58 | Yes |
| L80P | TM2 | 9.09 | 13 | 7.86 | 1.65 | 12.95 | Yes |
| L82P | TM2 | 9.78 | 26 | 7.05 | 3.69 | 9.62 | Yes |
| A106V | TM3 | 5.57 | 26 | 3.17 | 8.20 | 14.78 | No |
| A113P | TM3 | 7.60 | 12 | 10.80 | 1.11 | 5.51 | No |
| T118M | TM3 |  | 62 | 3.62 | 17.15 | 1.91 | Yes |
| S131C | Extracellular | 3.89 | 54 | 1.43 | 37.88 | -4.56 | Yes |
| A135T | TM4 |  | 83 | 1.58 | 52.53 | 2.88 | No |
| W140R | TM4 | 9.59 | 14 | 8.37 | 1.67 | 8.75 | Yes |
| L147R | TM4 | 7.36 | 7 | 2.16 | 3.24 | 13.74 | Yes |
| G150D | TM4 |  | 8 | 4.29 | 1.87 | 9.48 | No |
| G150C* | TM4 | 8.01 | 15 | 1.82 | 8.24 | 9.48 | No |
| R150H | Intracellular | 3.21 | 117 | 1.16 | 101.02 | -7.09 | Yes |
| WT |  |  | 100 | 1.00 | 100 |  |  |

**Supplementary Table 3.**
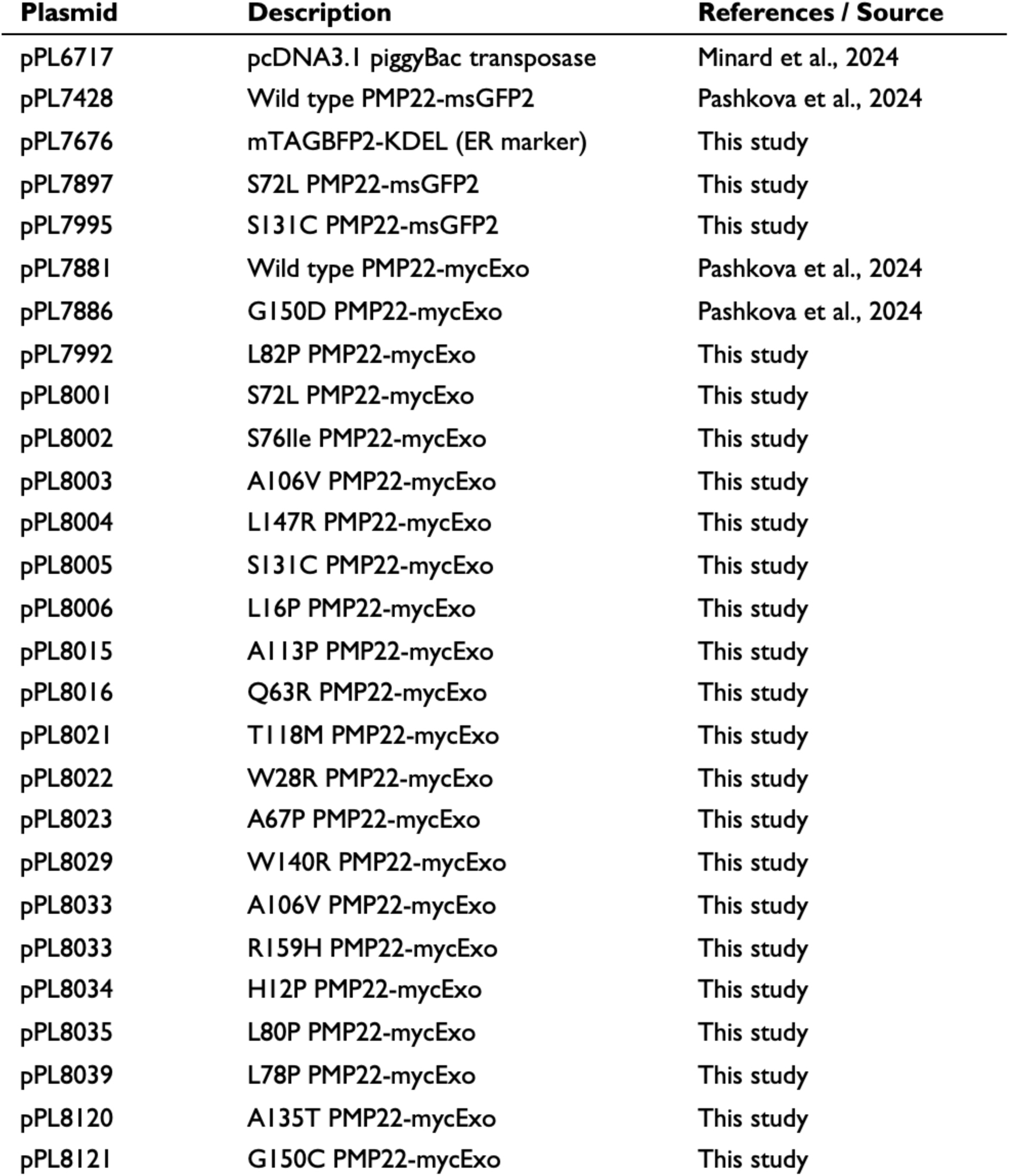
Plasmid DNA used in this study.

